# Virtual and remote opioid poisoning education and naloxone distribution programs: a scoping review

**DOI:** 10.1101/2023.11.15.23298586

**Authors:** Bruna dos Santos, Rifat Farzan Nipun, Anna Maria Subic, Alexandra Kubica, Nick Rondinelli, Don Marentette, Joanna Muise, Kevin Paes, Meghan Riley, Samiya Bhuiya, Jeannene Crosby, Keely McBride, Joe Salter, Aaron M. Orkin

**Author notes:** These authors contributed equally to this work (Bruna dos Santos, Rifat Farzan Nipun).

## Abstract

The opioid poisoning crisis is a complex and multi-faceted global epidemic with far-reaching public health effects. Opioid Poisoning Education and Naloxone Distribution (OPEND) programs destigmatize and legitimize harm reduction measures while increasing participants’ ability to administer naloxone and other life-saving interventions in opioid poisoning emergencies. While virtual OPEND programs existed prior to the COVID-19 pandemic and were shown to be effective in improving knowledge of opioid poisoning response, they were not widely implemented and evaluated. The COVID-19 pandemic brought both urgent and sustained interest in virtual health services, including harm reduction interventions and OPEND programs.

We aimed to assess the scope of literature related to fully virtual OPEND programming, with or without naloxone distribution, worldwide. A search of the literature was conducted and yielded 7,722 articles, of which 31 studies fit the inclusion criteria. Type and content of the educational component, duration of training, scales used, and key findings were extracted and synthesized. Our search shows that virtual and remote OPEND programs appear effective in increasing knowledge, confidence, and preparedness to respond to opioid poisoning events while improving stigma regarding people who use substances. This effect is shown to be true in a wide variety of populations but is significantly relevant when focused on laypersons. Interventions ranged from the use of videos, websites, telephone calls, and virtual reality simulations. A lack of consensus was found regarding the duration of the activity and the scales used to measure its effectiveness. Despite increasing efforts, access remains an issue, with most interventions addressing White people in urban areas. These findings provide insights for planning, implementation, and evaluation of future virtual and remote OPEND programs.

**Author Summary:** Facing a global health challenge, the opioid poisoning crisis affects individuals across all communities, ages, and socioeconomic groups, leading to high fatality rates. Educational programs addressing opioid poisoning have emerged as life-saving and cost-effective interventions. This review focuses on these programs conducted in a virtual setting, eliminating the need for in-person contact between staff and participants. We have identified and summarized evidence about the outcomes of these programs, which may include naloxone distribution. Our findings offer valuable insights for planning, implementing, and evaluating such programs. Furthermore, we highlight gaps in current knowledge, paving the way for future research.

## Introduction

The opioid poisoning crisis is a global health challenge with high fatality rates. The highest rates of opioid poisoning fatality are seen in the United States, followed by Estonia, Canada, and Lithuania(1–3). Opioid-related fatalities have been exacerbated by the increasingly toxic and unregulated drug supply market, contaminated by fentanyl and other substances such as benzodiazepines(2,4). Non-fatal opioid poisonings occur at an even higher rate than fatal opioid poisonings, adding to the social, health, and economic costs of the global opioid poisoning epidemic(5). Opioid-related harms can affect people in all communities, ages, and socioeconomic groups, including family members, friends, healthcare professionals and community members of people with lived or living experience (PWLLE) of opioid use(6,7).

Opioid Poisoning Education and Naloxone Distribution (OPEND) programs train and equip people to provide life-saving interventions, including naloxone administration(8–11). OPEND has been shown to save lives, improve knowledge and attitudes, and reduce stigma(8–13). However, rigorously evaluated OPEND programs have generally been limited to interventions involving in-person training and distribution, which can limit access to rural and remote populations and others who are unable or unwilling to access in-person services(14,15). As numerous programs pivoted away from in-person OPEND offerings during the COVID-19 pandemic, it brought a strong and sustained interest in the effectiveness and implementation of virtual OPEND programming(16).

This scoping review aims to assess the range of literature concerning opioid poisoning education programming conducted entirely without in-person interaction. In compiling a cohesive overview of existing virtual OPEND programs reported in the literature, we aim to support the development and evaluation of future programs.

## Methods

### Objective and Review Question

We synthesized the literature concerning virtual OPEND programming for people who may witness an opioid poisoning. We structured our scoping review question based on the Participants, Concept, Context framework for scoping reviews(17).

- Participants: People at risk of opioid poisoning or likely to witness opioid poisoning or otherwise interested in OPEND program participation.
- Concept: Any opioid poisoning education programming with or without naloxone distribution that is conducted entirely at a distance, without in-person interaction between participants and program personnel.
- Context: Worldwide.

### Language and Terminology

Although ‘overdose’ is a common term, we use ‘opioid poisoning’ throughout this paper. ‘Overdose’ suggests that the primary cause for the health emergency is a matter of incorrect dose, which can place stigmatizing blame on people who experience opioid poisoning. ‘Poisoning’ draws attention to the broader social context leading to these emergencies and the epidemic of toxic drug harms. We also prefer the term ‘opioid poisoning education and naloxone distribution’ over ‘take-home naloxone’ because the latter does not fully acknowledge the circumstances of the many people who experience opioid poisoning and homelessness and does not sufficiently acknowledge the educational and non-pharmacological elements of these programs.

### Approach and Protocol

The design and methods for this review are reported in accordance with the Preferred Reporting Items for Systematic Reviews and Meta-Analyses Extension for Scoping Reviews (PRISMA-ScR)(18). Further guidance was gained from an adapted version of Arksey and O’Malley’s methodological framework on scoping reviews(19) and the Joanna Briggs Institute methodological guidance for conducting systematic scoping reviews(20). A search of MEDLINE and the Cochrane Database of Systematic Reviews was conducted and no current or underway literature reviews on the topic were identified. We developed a review protocol and registered it on Open Science Framework (osf.io/ew9rq)(21).

### Search Strategy

An initial limited search on MEDLINE was undertaken to identify papers on the topic using terms according to the Participants, Concept, Context framework(17), including the terms ‘virtual’, ‘education’, ‘opioids’, and related terminology. A University of Toronto health science librarian provided expertise in optimizing the search strategy according to project objectives. The text words contained in the titles and abstracts of relevant papers and index terms were used to develop a full search strategy for OVID MEDLINE, OVID Embase, EBSCO CINAHL, OVID PsycINFO, The Cochrane Library, SCOPUS, and ERIC (S1 Table). The search strategy, including all identified keywords and index terms, was adapted for each included database. Scholarly database searches were conducted on June 1, 2023. The grey literature search was conducted on July 27, 2023, including the Canadian Agency for Drugs and Technologies in Health’s (CADTH) Grey Matters website, the System for Information on Grey Literature in Europe (OpenGrey), and TRIP Pro. We screened the references of included papers to identify additional papers. All supplementary searches are documented in S1 Table.

### Source of Evidence Selection

Following deduplication, titles and abstracts were screened independently by four reviewers (BDS, RFN, AK, and AMS) based on the inclusion criteria. The full-text review of the selected citations was undertaken by BDS and RFN. Reasons for exclusion during full text review were recorded in a PRISMA flow diagram (Fig 1)(18). Any disagreements between the reviewers at any stage of the selection process were resolved through discussion and consensus.

**Figure 1.**
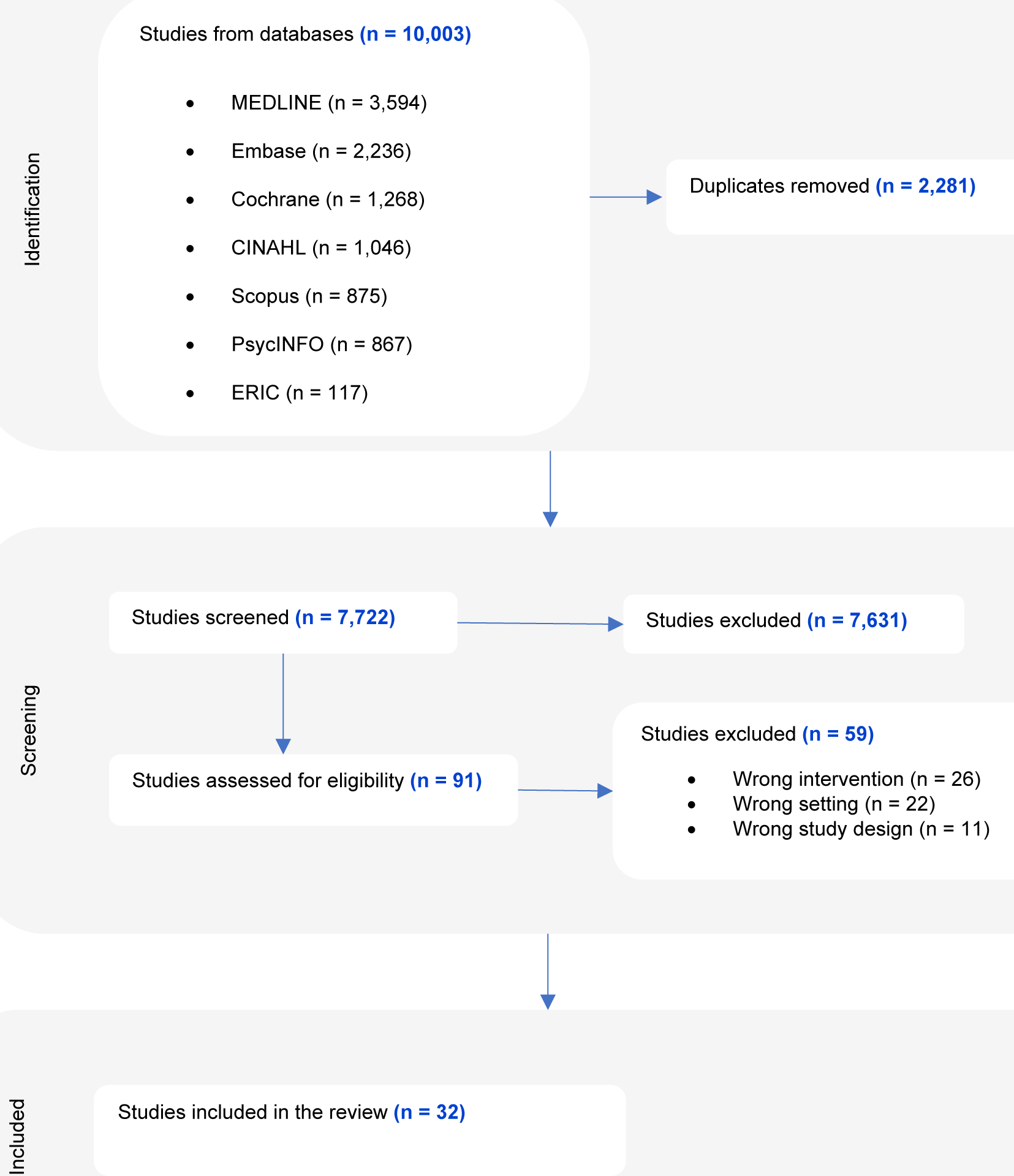
PRISMA flowchart. This figure depicts studies that were included in the scoping review at each stage of the review process.

### Data Extraction

Data were extracted in Covidence from included sources by two independent reviewers (BDS and RFN) using a data extraction tool developed by the reviewers and modified from the JBI Manual for Evidence Synthesis(22). The data extracted included publication details (year of publication, country, and type of paper), information about the target population (demographics such as age, gender, and race/ethnicity), intervention components (e.g., internet-based, telephone-based), educational components (topics addressed by the educational portion), key findings, and other details relevant to the review question. All variables extracted are provided (S2 Table). The data extraction tool was first piloted and modified as necessary with ten papers divided between the two reviewers. Any disagreements between the reviewers were resolved through discussion and consensus.

### Data Synthesis

The extracted data are synthesized and summarized in narrative and tabular format. When appropriate, findings were grouped based on themes.

## Results

After deduplication, 7,722 out of the initial 10,003 papers were identified for further review. After title and abstract screening, 7,631 papers were excluded, and 91 were reviewed in full text (Fig 1). Full text review elicited 32 papers for inclusion within this scoping review. Sisson et al., 2023 conducted a single study that was reported in two papers and were, therefore, treated and reported here as a single study(23,24).

Characteristics of included papers are specified in Table 1. Seventeen studies have a quasi-experimental pre-post design, eleven are descriptive, two are randomized controlled trials, and one is a non-randomized controlled trial. The publication period extended from 2016 to 2023, peaking notably in 2022 (25%, n=8). Of the studies included in our analysis, 30 (97%) were based in the United States of America; one was conducted in Australia.

**Table 1.**
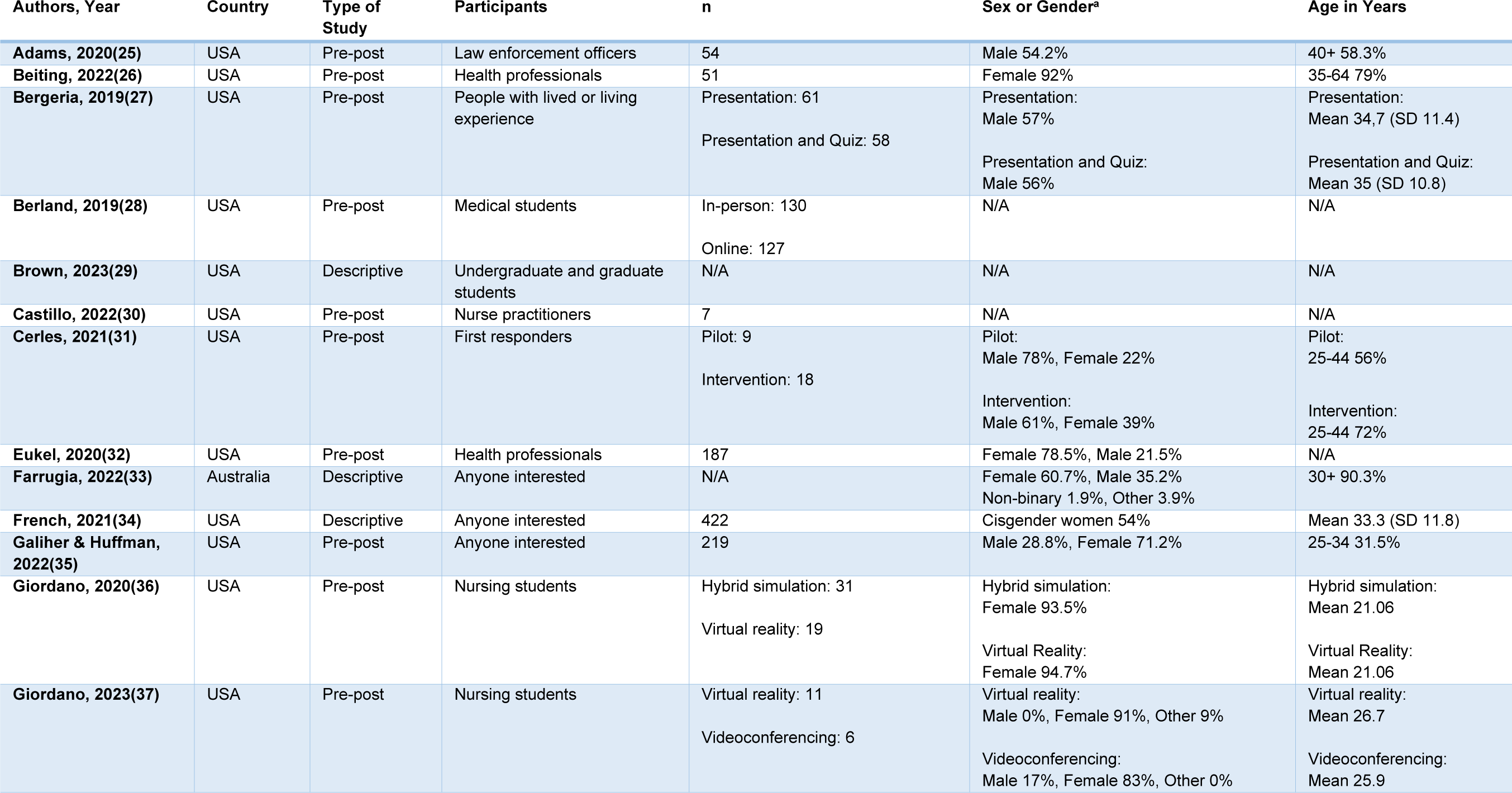
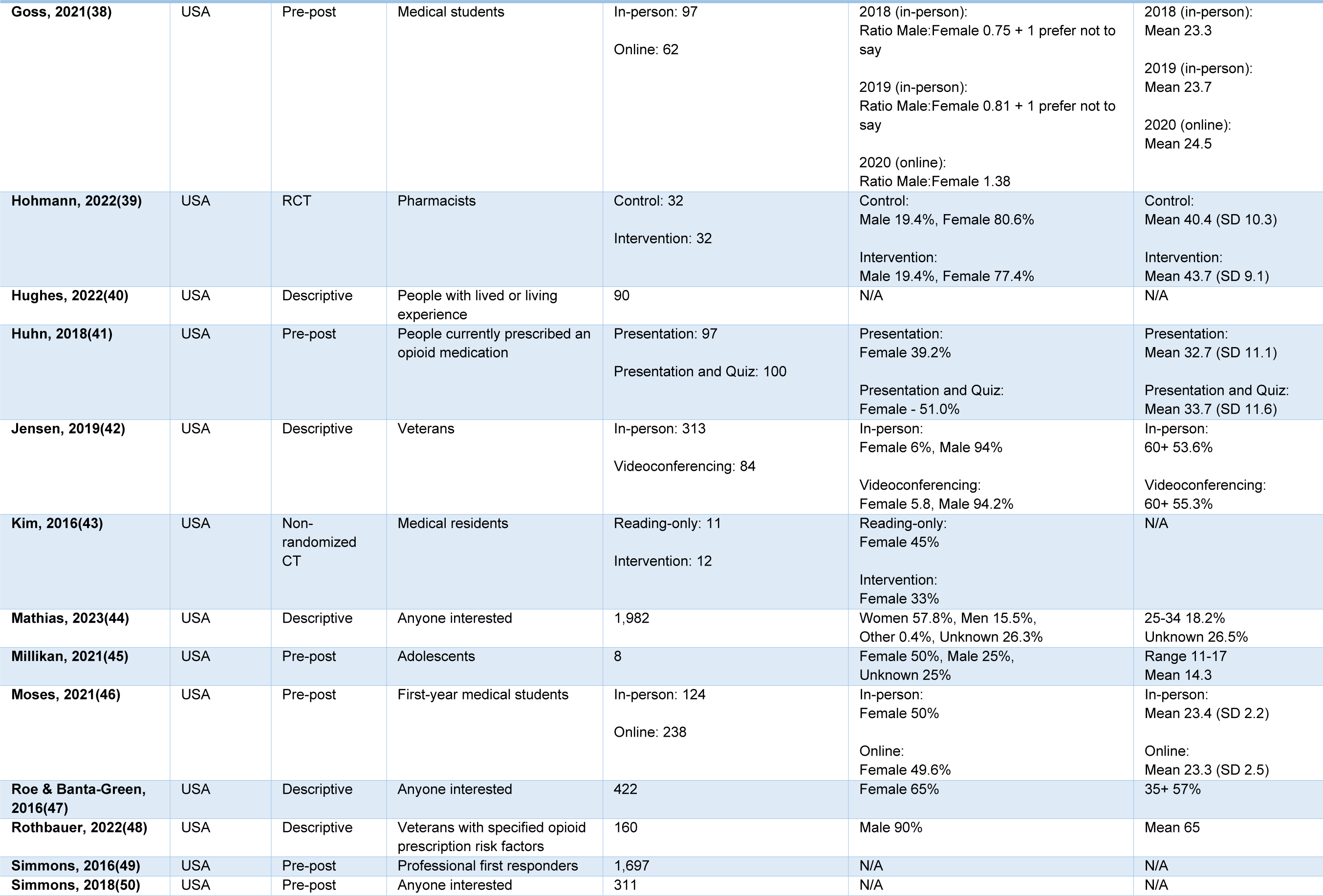
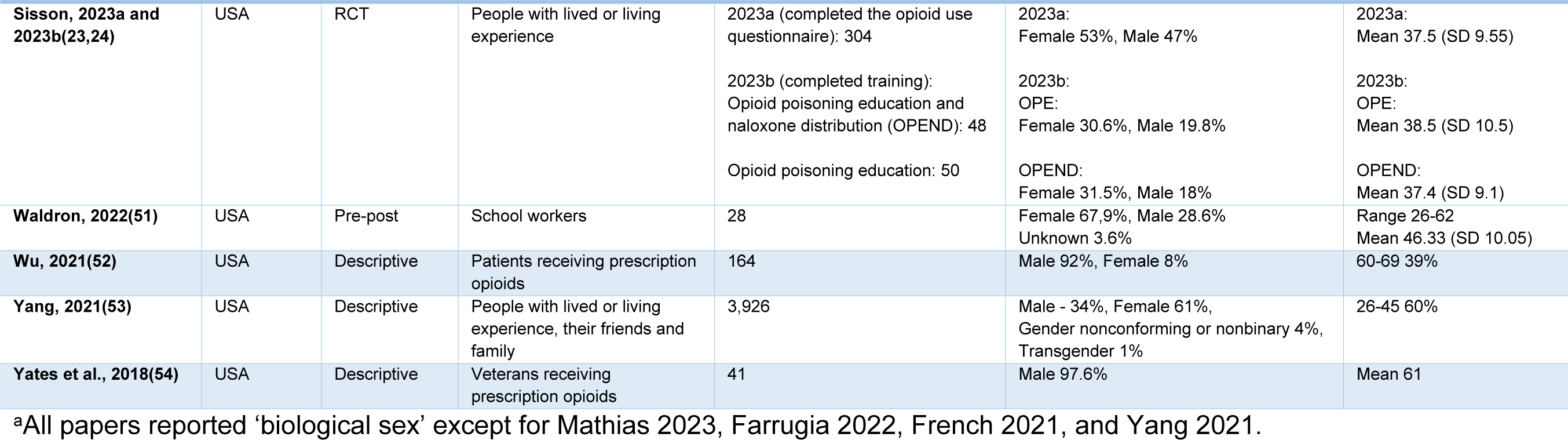
Characteristics of included papers. This table shows the characteristics of papers included in this scoping review.

### Participant Demographics

Eight of the included studies (26%) referred to interventions that were designed specifically for people with lived or living experiences (PWLLE) of opioid use, six (19%) broadened to anyone interested in participating, including laypersons, and other six (19%) focused on medical, nursing and pharmacy students. The remainder of the studies involved health professionals (n=5), family members or friends of PWLLE (n=2), first responders (n=2), school workers (n=1), and law enforcement officers (n=1).

Of the 31 studies analyzed, fourteen (45%) reported on the mean age of the participants. The ages ranged from 11 to 69 years old, with most of the participants aged between 25 and 64 years. Of the included studies, 21 (68%) reported biological sex and not gender, with ten of these which reported male and female options. There was notable variation in participation based on biological sex, with thirteen studies (42%) primarily comprised of female participants, and nine (29%) of male participants. Four studies (13%) included an option for participants to report their gender, while six (19%) did not report any sex or gender information.

Data on race or ethnicity were not reported in thirteen (42%) of the studies. Three studies (10%) reported the percentage of one racial identity in their results, with Beiting et al, 2022 reporting the percentage of Black individuals, while Yates et al., 2018 and Moses et al., 2021 reported the percentage of White individuals. It is worth noting that all of the 18 studies reporting race or ethnicity identity, and had White or Caucasian participants as the majority(23,25,26,31,34–37,39,42,44–46,52–55).

No papers provided information on participants’ socioeconomic status. However, Huhn et al., 2018 reported that the median household income of participants was US$52,500, while Sisson 2023a reported that most participants had an annual household income between US$10,000 and US$49,000. Participants educational level was available in six (19%) studies, with three reporting most participants had a high school degree(23,27,55), two reporting most had a college degree(35,51), and two reporting most had a graduate degree or higher(32,39). The geographical area in which the participants resided was mentioned in six studies. Most participants were from urban areas(28,36,37,39), while two studies focused on rural areas(30,51).

### Intervention Approaches

Several strategies were employed for conducting remote and virtual OPEND. A summary of intervention approaches is specified in Table 2. A significant number of these studies (n=25, 81%) delivered interventions over the internet, including videos (n=9), videoconferencing (n=7), courses (n=5), slide presentations (n=3), and content on websites (n=3). Of these, four studies additionally provided naloxone by mail. There were four telephone-based interventions, all of which were coupled with mail-delivered naloxone.

**Table 2.**
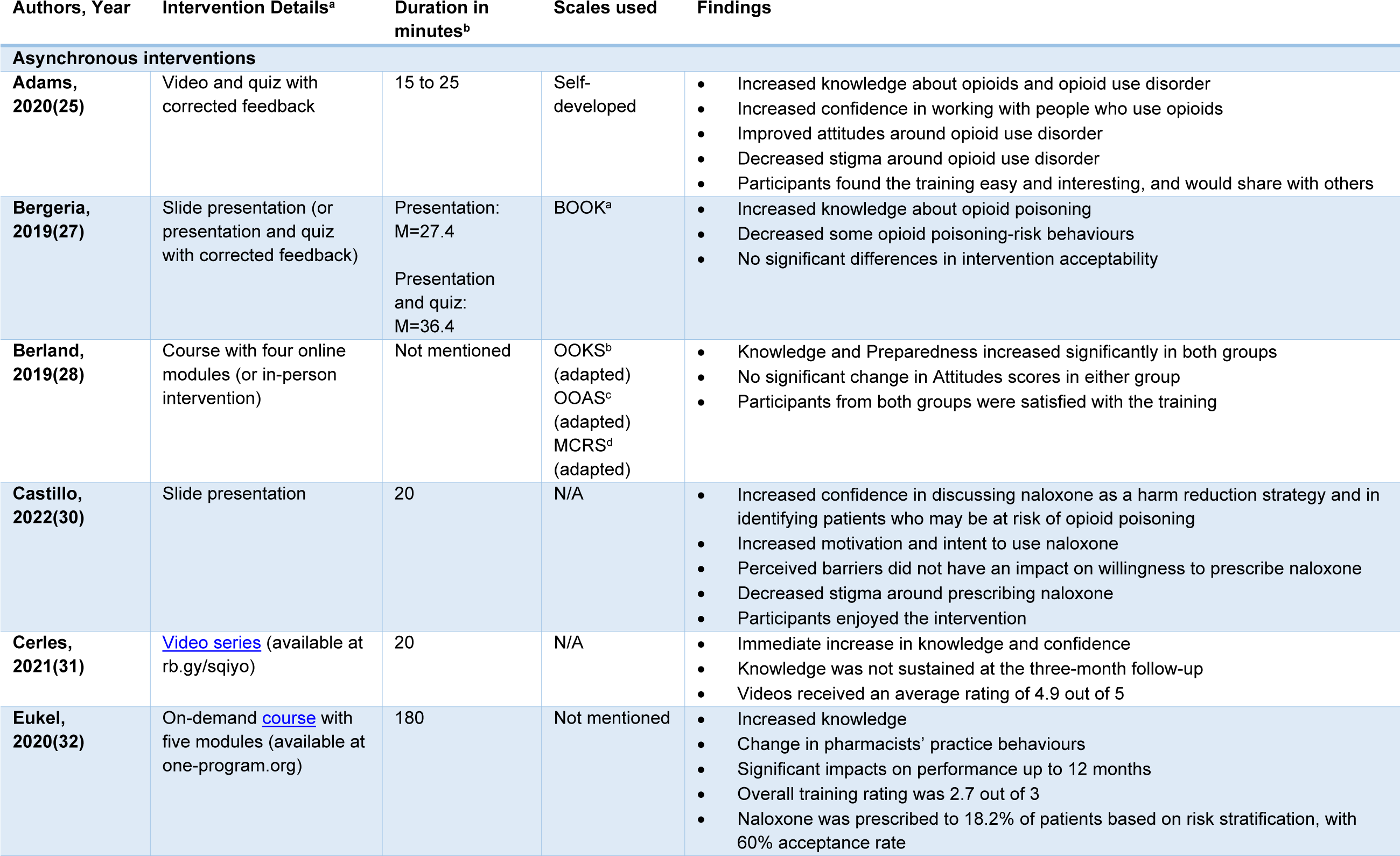
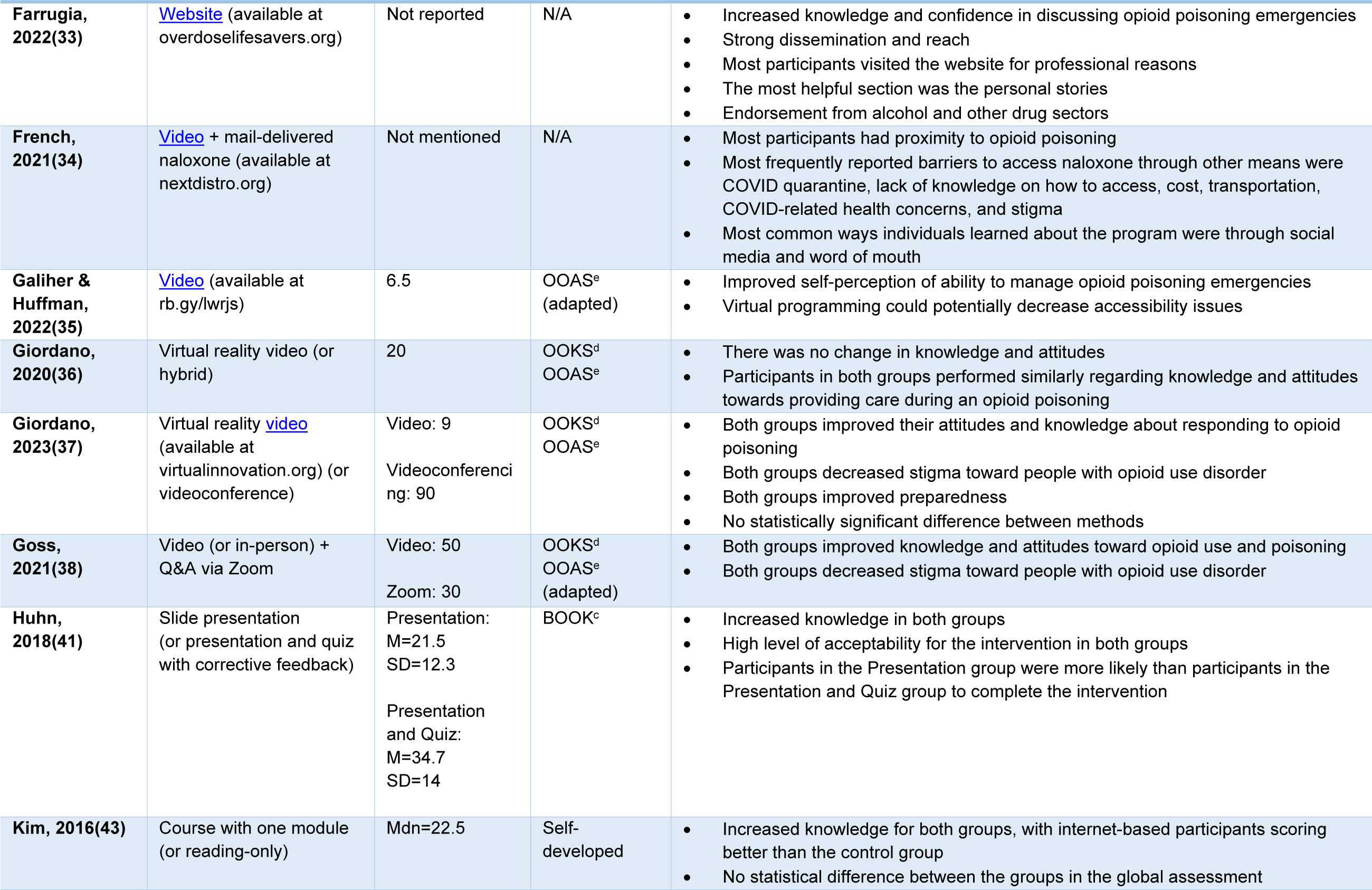
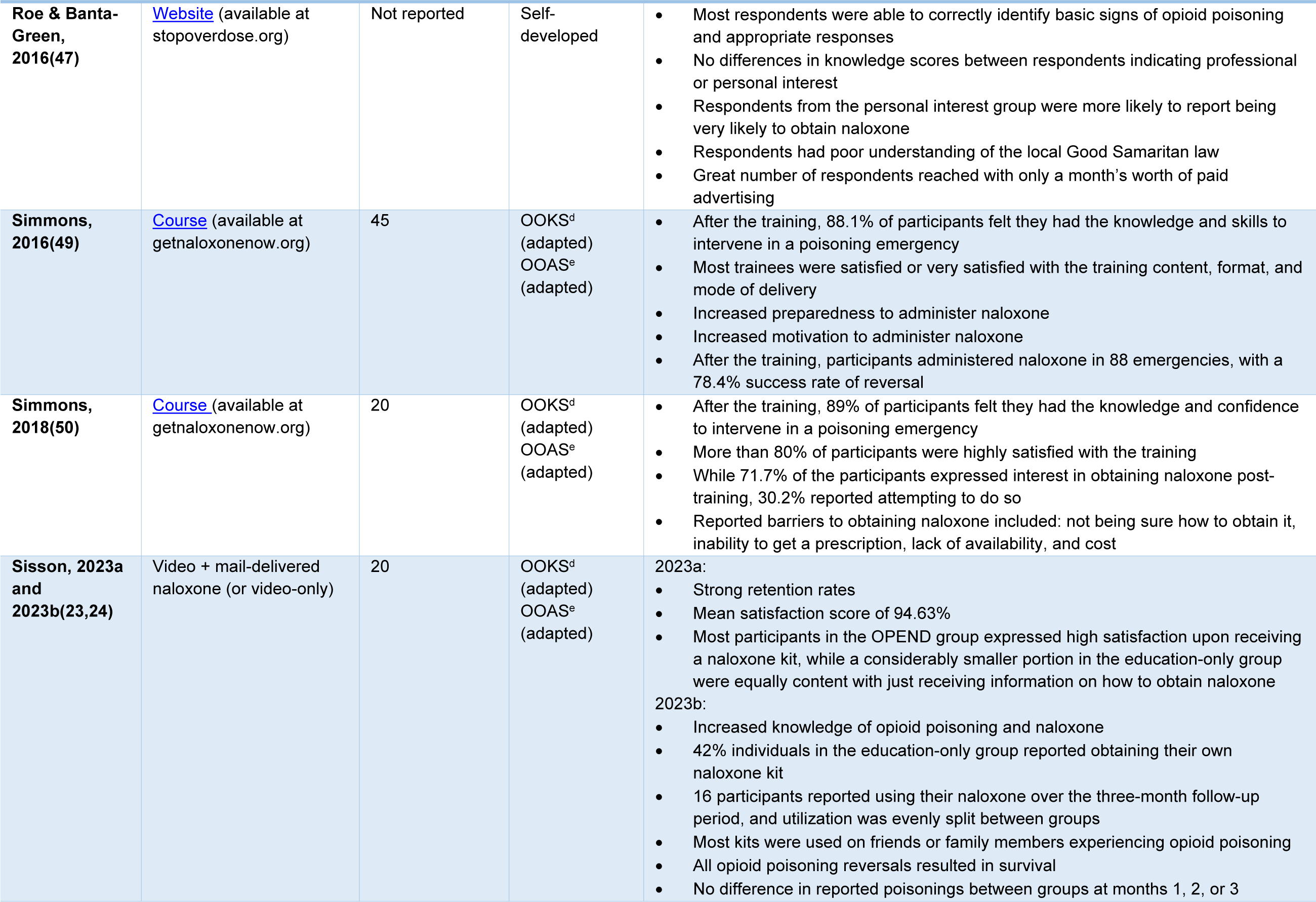
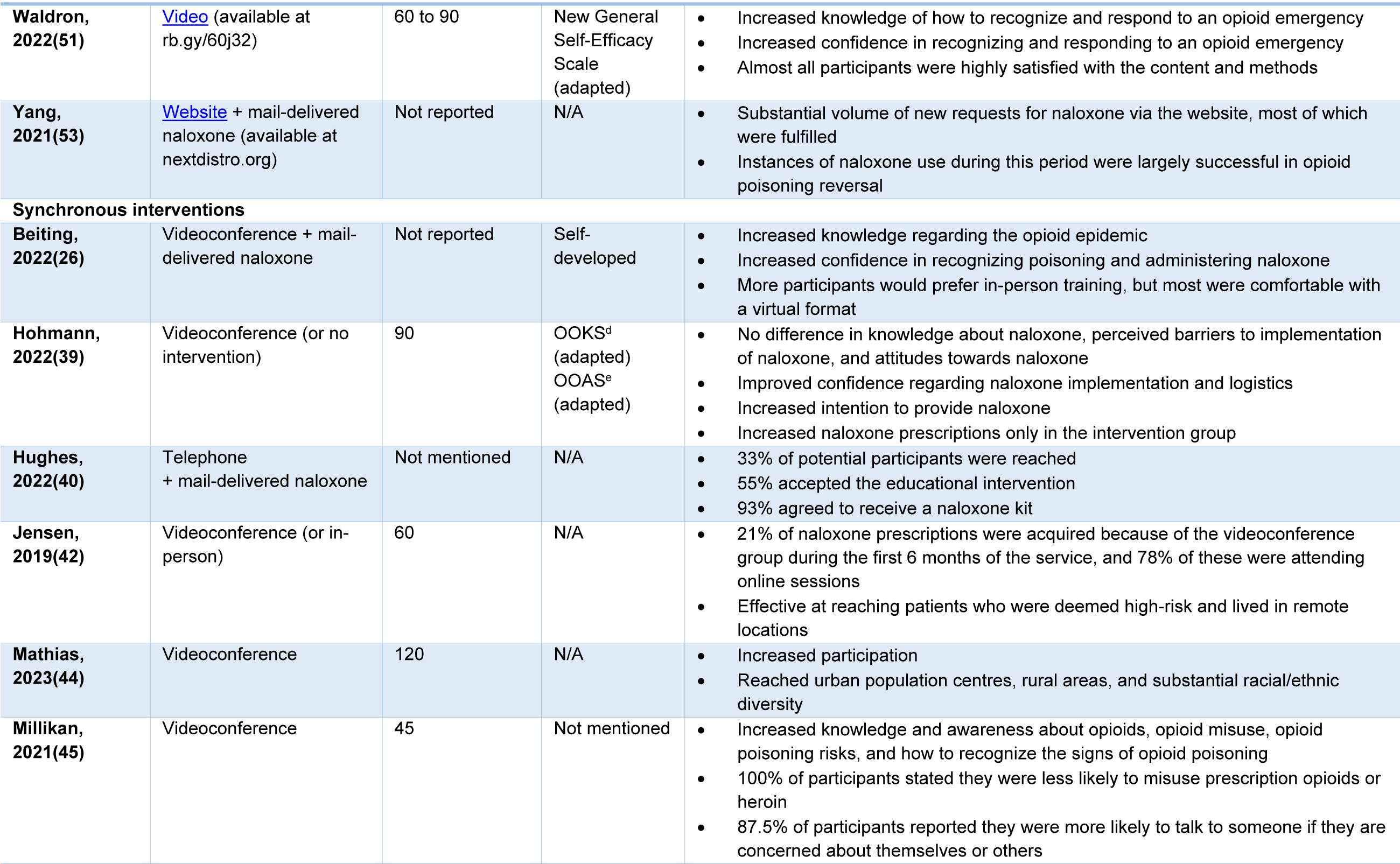
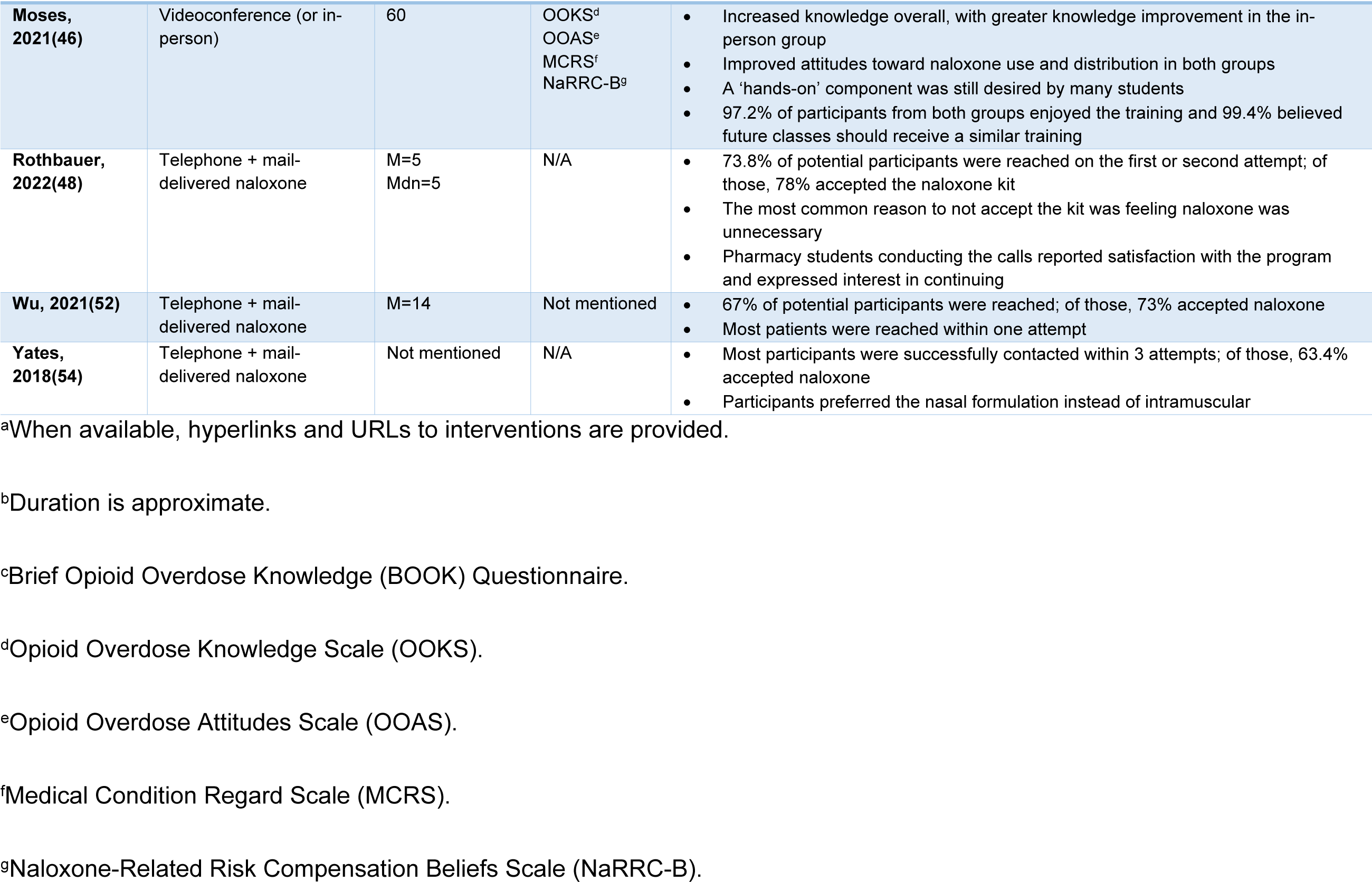
Intervention characteristics and key findings. This table shows details and key findings of the interventions addressed in this review.

Among the 27 studies that reported on the duration of the intervention, the educational components ranged from five minutes to two hours, with interventions typically lasting 20 minutes (n=6, 19%). Four studies (13%) used a self-directed approach, in which participants could complete the intervention in their own time(30,32,33,47). Of the self-directed interventions, one reported the average amount of time taken for participants to complete their training, and none reported drop-out rates.

The 31 studies analyzed in this review incorporated a variety of educational elements in their interventions. The predominant theme among the educational components analyzed was how to respond to, followed by how to recognize, and how to prevent opioid poisoning emergencies. Typical opioid poisoning response steps were sporadically mentioned across different interventions, with the most often mentioned being naloxone administration and calling 911. Other themes found within the interventions included history and statistics on opioid poisoning, laws and law enforcement, specifics on naloxone use, and stigma. An overview of the educational components can be seen in Table 3.

**Table 3.** Thematic Analysis of Educational Components. This table shows a thematic analysis of the topics mentioned in the educational portion of the interventions.

| Major Themes<sup>a</sup> | Subthemes | References |
| --- | --- | --- |
| Statistics and History of Opioid Poisoning |  | 8 |
| Prevention of Opioid Poisoning | Opioid poisoning risks | 8 |
|  | Contamination by fentanyl and/or other substances | 2 |
| Recognizing an Opioid Poisoning |  | 19 |
| Responding to an Opioid Poisoning | PPE and situational awareness | 2 |
|  | Calling 911 | 7 |
|  | Airway and circulation assessment | 1 |
|  | Sternal rub | 1 |
|  | Naloxone administration | 22 |
|  | Rescue breathing | 3 |
|  | Recovery positioning | 4 |
|  | Opioid withdrawal management | 3 |
|  | Continued monitoring until EMS arrives | 3 |
| **Laws and Law Enforcement** | Barriers to calling 911 | 2 |
|  | Good Samaritan Law | 5 |
|  | Naloxone Access Law | 2 |
| **Naloxone Specifics** | Intranasal vs. intramuscular use | 1 |
|  | Mechanism of action | 2 |
|  | Access and availability | 1 |
|  | Storage and expiration | 1 |
| **Stigma about Opioid Poisoning** |  | 6 |

<sup>a</sup>Themes were identified based on information reported in papers, or through direct revision of original educational materials, when available.

Most studies utilized a pre-post design or post-training questionnaires. Out of the 31 studies analyzed, 19 (61%) reported on the level of knowledge individuals possesssed regarding opioid poisoning response, followed by 12 (39%) on attitudes and behaviour towards opioid poisoning emergencies, 11 (36%) on qualitative feedback about the intervention, and 10 (33%) on the number of naloxone kits that were distributed, prescribed, or obtained during the intervention.

Other outcomes assessed were confidence in responding to an opioid poisoning emergency, comparisons between different modalities, accessibility of OPEND, changes in stigma, barriers to accessing or prescribing naloxone, preparedness, familiarity with virtual learning, and opioid use.

The Opioid Overdose Knowledge Scale (OOKS) and the Opioid Overdose Attitudes Scale (OOAS) were used in 10 out of 31 studies (32%). Eight studies used both scales, while two used either one of them. Three studies developed their own assessment tools, some drawing from various sources in the literature. The Brief Opioid Overdose Knowledge Questionnaire (BOOK) and the Medical Conditions Regard Scale (MCRS) were each used by two studies (6%).

## Key Findings

Findings are summarized in Table 2.

### Videos (n=9)

Six studies reported an increase in knowledge immediately after the training(24,25,31,37,38,51). One of these collected longitudinal data, and knowledge was not sustained at the three-month follow-up(31). Three of the video interventions reported on attitudes around opioid use disorder (OUD) or providing care during an opioid poisoning(25,36,37), with one of them reporting no change(36) and the other two reporting an increase in positive attitudes. All four studies that assessed overall satisfaction and/or qualitative feedback reported high satisfaction rates regarding the content, teaching methods, format, and/or mode of delivery of the intervention (23,25,31,51). Other reported outcomes were increased confidence, self-perception of ability to manage opioid poisoning emergencies, and preparedness to recognize and respond to opioid poisoning, and decreased stigma around OUD.

### Videoconferencing (n=7)

Four studies reported an increase in knowledge about the use of naloxone and the opioid poisoning epidemic(26,37,45,46), and one reported no difference(39). Two papers reported improved attitudes toward naloxone use and distribution(37,46), while another reported no difference after the training(39). Two papers reported that most participants are comfortable with a virtual format but would prefer in-person training(26,46).

### Courses (n=5)

All studies reported increased knowledge and awareness about opioid poisoning(28,32,43,49,50). While one study reported a positive change in behaviour after the intervention(32), another study found no significant changes for attitudes pertaining to opioid poisoning, as measured using the validated OOAS tool(28). Four papers that assessed satisfaction with the training reported high satisfaction rates(28,32,49,50). Simmons et al., 2016 reported that participants surveyed up to 9 months after the intervention successfully reversed an opioid poisoning with the use of naloxone in 78.4% of cases(49).

### Slide presentations (n=3)

All studies reported high levels of satisfaction and acceptability for their interventions(27,30,41). Two studies reported increased knowledge and one increased confidence about opioid use and poisoning after the intervention(27,30,41). One study found that the training was effective in decreasing stigma around prescribing naloxone(30). One study reported less opioid poisoning-risk behaviours after the intervention(27).

### Websites (n=3)

The types of assessments used in website interventions varied significantly. One study reported increased knowledge and confidence in discussing opioid poisoning(33), and another showed that most participants correctly identified basic signs of opioid poisoning and appropriate responses(47). One of the interventions reported that of the participants visiting the website, a majority indicated that they visited for professional reasons(33). Another intervention showed no difference in knowledge between participants that indicated professional or personal interest, with the personal interest group were more likely to obtain naloxone(47).

### Telephone (n=4)

All telephone-based interventions reported the percentage of participants successfully contacted through the intervention. In three of them, more than half were reached within one or two attempts(48,52,54).

### Mail-delivered naloxone (n=8)

Of the interventions that offered naloxone by mail, four were telephone-based(40,48,52,54), two used videos(23,24,34), one used videoconference(26), and one used a website(53). Four studies reported acceptability rates for naloxone kits ranging from 73% to 93%(40,48,52,54). Among participants that did not accept naloxone, one study reported the most common reason was feeling it was unnecessary(48). One study reported on barriers to accessing naloxone through means other than the intervention, with the most frequently reported barriers being the COVID-19 epidemic, lack of knowledge on how to access naloxone, cost, transportation, and stigma(34). Two papers reported the result of participants’ naloxone use – both papers reported that most or all opioid poisoning reversals resulted in survival, and one paper reported that a majority of kits were used on friends or family members(24,53).

### Comparison between different modalities (n=12)

Table 2 indicates which studies had comparison groups. Four studies had a control group involving in-person training(28,38,42,46), and one study had a hybrid group(36). Of these, three showed no significant differences on preparedness, attitudes, satisfaction, and stigma scores between formats(28,36,38), and one reported greater knowledge in the in-person group(46). One study reported a significantly higher number of naloxone prescriptions obtained by participants in the virtual group(42). For papers with other comparison groups, there was no significant differences in Kim et al., 2016 between reading-only and course groups, in Bergeria et al., 2019 for slide presentation and slide plus quiz groups, and in Giordano et al., 2023 between virtual reality video and videoconference groups(27,37,43). Huhn et al., 2018 showed the presentation-only group were more likely to complete the intervention compared to the group that received the same presentation coupled with a quiz, but there were no differences in knowledge and intervention acceptability(41). Sisson et al., 2023a reported higher satisfaction in the naloxone distribution group compared to the group without naloxone distribution(55). Sisson et al, 2023b reported that 42% of individuals in the group without naloxone distribution obtained their own kit(24). In a comparison between a videoconference intervention and no intervention, Hohmann at al., 2022 showed an increase in the number of naloxone prescriptions obtained by participants only in the intervention group, but there was no difference related to other outcomes(39).

### Limitations

The included studies reported on limitations in their designs. Commonly reported limitations included lack of generalizability of their study results, reported in 21 studies(23–31,36–39,44– 50,52,54). A frequent obstacle reported in 12 studies is small sample size, which ranged from six to 313 participants per group(25–27,30,31,36,37,42,45,48,51). Other limitations include the absence of longitudinal data(26,34,39,41,45,47,52), self-reported bias for questionnaires(23,24,46,49,50), the lack of comparison to an in-person intervention(35,38,41,44), no follow-up on the number of participants who responded to an opioid poisoning post-intervention(34,42,44,52), lack of validity of the tools used(25,30,32), low response rates, which were of 21.2% and 33.7%(32,50), and the lack of randomization(43,44).

## Discussion

This scoping review identified that there is considerable research and innovation underway in the development and evaluation of virtual OPEND, particularly since the start of the COVID-19 pandemic. Overall, virtual OPEND appears to be acceptable to participants and effective in increasing knowledge on how to respond to an opioid poisoning, as well as improving attitudes and decreasing stigma about people who use opioids. Especially when coupled with naloxone distribution, programs appear to impact the motivation and willingness of people to respond to these emergencies. Two studies demonstrated successful resuscitations and decreased poisoning death rates(23,24,49), showing that virtual OPEND can be a life-saving intervention.

Our analysis indicates that most interventions are primarily targeted at PWLLE of opioid use and potential bystanders, defined as individuals who could potentially witness an opioid poisoning. A cross-sectional web-based survey and a mixed-methods study have demonstrated the unique position of bystanders in reversing opioid poisonings and saving lives, provided they are equipped with naloxone and the knowledge to administer it effectively(56,57).

However, the bulk of these studies are based in the USA, with only one study conducted in Australia(33). Opioid poisoning is a global concern, demonstrating a pressing need to implement and evaluate virtual OPEND programs worldwide to increase access to this intervention(2). There is also a need for more randomized trials, since only two included studies used this design. Few studies had a longitudinal design; however, it is essential to assess the attrition of knowledge acquired in these interventions and the potential need for retraining.

In addition, only 42% of studies utilized validated scales to measure outcomes related to opioid poisoning, and 62% of those studies modified already validated tools for their study. The validated scales were originally designed to assess illicit opioid poisoning risk factors and do not include knowledge about naloxone(58,59). Therefore, there is a critical demand for updated and validated survey instruments to effectively evaluate these interventions(58,59).

Few studies reported sociodemographic variables and focused on the inclusion of historically marginalized populations. Yet, those more likely to be impacted by the opioid poisoning crisis include people living in rural and remote areas, individuals experiencing homelessness, those living in poverty, incarcerated individuals, and Black, Indigenous, and People of Colour(60). Without sociodemographic information, it is unknown whether sampling and selection biases and issues with representativeness have occurred, limiting the effectiveness of these interventions in addressing health inequities(61).

Most interventions did not make their training materials publicly available, and some of them did not specify the educational content included in the intervention, hindering the potential for public dissemination and research replication. Among the studies that reported the content of their training, stigma reduction and policies pertaining to witness protection and naloxone permission for opioid poisoning reversal were frequently not addressed, which if included, could have bolstered the outcomes(62,63).

This is the first scoping review on the outcomes of fully virtual OPEND interventions. Other reviews have explored opioid poisoning interventions and their effectiveness. Pellegrino et al., 2021 has conducted a scoping review on first aid educational interventions for opioid poisoning published until 2019, showing the potential of these programs despite the sparsity and lack of quality of the studies reviewed(11). Razaghizad et al., 2021, an umbrella review on OPEND, demonstrated that there is credible evidence that opioid poisoning education programs improve knowledge and attitudes, enable participants to use naloxone safely and effectively, and reduce opioid-related mortality(10). Additionally, Tas et al., 2022 reviewed technological interventions that prevent, detect, or respond to opioid poisoning emergencies, with promising approaches that were still under development(64). Our findings suggest the potential effectiveness of virtual OPEND interventions and can assist researchers and public health practitioners to design, implement, and evaluate future virtual and remote OPEND programs. Moreover, our findings suggest the urgent need for more rigorous research of virtual OPEND programs, which have the potential to save lives. Our recommendation includes providing resources and conditions to amplify these educational programs coupled with naloxone distribution as a health care tool to reduce opioid-related harms and inequities worldwide. Future research is recommended, especially in developing relevant evaluation tools that can better assess OPEND’s effectiveness for diverse outcomes. Longitudinal clinical trials are also encouraged to address the attrition of knowledge and more controlled settings.

## Limitations

Our review has limitations. One such limitation is the lack of extensive critical appraisal of the studies included. Despite our best efforts to design a comprehensive search strategy without imposing any limitations, the constant updates and rapid publication of new research on technology-based interventions may result in the unintentional omission of significant studies. We also acknowledge that many impactful programs addressing the opioid poisoning crisis are currently available online but did not involve a research design or share publicly available data on these interventions, therefore were not suitable for this review. Additionally, the lack of detailed information about the interventions in many studies may have impacted the accuracy of our synthesis, as we did not reach out to the authors for additional information.

## Conclusion

This review synthesizes the literature on virtual and remote Opioid Poisoning Education and Naloxone Distribution (OPEND) programs, and underscores gaps in program implementation and evaluation. Currently, it is difficult to draw substantial conclusions on OPEND effectiveness due to the variety of study designs and the lack of updated and validated evaluation methods. However, existing evidence points to the potential to increase knowledge, decrease stigma, and ultimately reduce morbidity and mortality due to opioid-related harms. This life-saving knowledge can significantly and positively impact populations who are not currently being adequately reached by in-person OPEND interventions.

## Authors’ Contributions

Conceptualization: AMS, AK, AO, NR, DM, JM, KP, MR, SB, JC, KM, JS

Methodology: AMS, AK, AO, BDS, RFN

Formal analysis: BDS, RFN

Investigation: BDS, RFN

Data curation: BDS, RFN

Writing – original draft: BDS, RFN

Writing – review and editing: AMS, AK, AO

Visualization: BDS, RFN

Supervision: AMS, AK, AO

Project administration: AMS, AK

Funding acquisition: AO, DM, JM, KP, MR, SB, JC, KM, JS

## Data Availability

All relevant data are within the manuscript and its Supporting Information files.

## Acknowledgements

We acknowledge the time and efforts of Eden Kinzel, Liaison and Education Librarian at the University of Toronto, who provided expertise in the development of the research question and search strategy.

## Supporting information

**S1 Appendix. Search strategy.**

**S2 Appendix: Data extraction instrument.**

